# Identifying common pharmacotherapies associated with reduced COVID-19 morbidity using electronic health records

**DOI:** 10.1101/2020.04.11.20061994

**Authors:** Victor M Castro, Rachel A Ross, Sean M McBride, Roy H Perlis

**Author notes:** **Correspondence:** Roy H. Perlis, MD MSc, Massachusetts General Hospital, 185 Cambridge Street, 6th Floor, Boston, MA 02114.

## Abstract

**Objective:** Absent a vaccine or any established treatments for the novel and highly infectious coronavirus-19 (COVID-19), rapid efforts to identify potential therapeutics are required. We sought to identify commonly prescribed medications that may be associated with lesser risk of morbidity with COVID-19 across 6 Eastern Massachusetts hospitals.

**Design:** In silico cohort using electronic health records from individuals evaluated in the emergency department between March 4, 2020 and July 12, 2020.

**Setting:** Emergency department and inpatient settings from 2 academic medical centers and 4 community hospitals.

**Participants:** All individuals presenting to an emergency department and undergoing COVID-19 testing.

**Main Outcome or Measure:** Inpatient hospitalization; documented requirement for mechanical ventilation.

**Results:** Among 7,360 individuals with COVID-19 positive test results by PCR, 3,693 (50.2%) were hospitalized in one of 6 hospitals. In models adjusted for sociodemographic features and overall burden of medical illness, medications significantly associated with diminished risk for hospitalization included ibuprofen and sumatriptan. Among individuals who were hospitalized, 962(26.0%) were admitted to the intensive care unit and 608 (16.5%) died; ibuprofen and naproxen were also more commonly prescribed among individuals not requiring intensive care.

**Conclusions:** These preliminary findings suggest that electronic health records may be applied to identify medications associated with lower risk of morbidity with COVID-19, but larger cohorts will be required to address the substantial problem of confounding by indication, such that extreme caution is warranted in interpreting nonrandomized results.

**Trial Registration:** None

**Summary Boxes:** *Section 1: What is already known on this topic:* Absent a vaccine or any established treatments for the novel and highly infectious coronavirus-19 (COVID-19), rapid efforts to identify potential therapeutics are required.

*Section 2: What this study adds:* This cohort study across 6 hospitals identified medications enriched among individuals positive for COVID-19 who are less likely to experience adverse outcomes including hospitalization, intensive care, or death.

## Introduction

The rapid spread and mortality associated with the 2019 novel coronavirus (2019-nCoV; severe acute respiratory syndrome coronavirus 2 (SARS-CoV-2), or COVID19) necessitates efforts to find therapeutic strategies with unprecedented speed. Absent any treatments known to be effective against the novel virus, randomized trials have been launched for antiviral agents already in development for other indications[1], and in vitro studies[2] or in silico modeling efforts [3] have identified compounds with potential antiviral benefit.

If medications already known to be safe and approved for human use exhibited benefit, they could rapidly be deployed in clinical settings once efficacy was established in randomized trials. Such repositioning has been embraced enthusiastically in other contexts [4]. For example, we previously demonstrated that one commonly-prescribed medication could shorten, rather than prolong, QT interval[5]. However, with COVID-19, preliminary efforts to reposition the antimalarial medications hydroxychloroquine or chloroquine demonstrated the speed with which enthusiasm far outstripped data [6].

In the present study, we aimed to use electronic health records (EHR) to identify medications that might be associated with decreased risk of COVID-19-associated morbidity, including hospitalization and intensive care unit (ICU) admission and death. In particular, we examined medications more commonly prescribed in the period prior to hospitalization to individuals with documented infection who did not experience such morbidity. While adequately powered and well-designed randomized trials are required to demonstrate efficacy, we hypothesized that a simple pharmacovigilance strategy examining enrichment of individual medications in the morbidity-free group would facilitate identification of candidate drugs for efficacy studies and prioritization of medications implicated by cellular screens.

## Methods

### Subjects

We included all individuals undergoing COVID-19 testing through one of 6 hospitals, including 2 academic medical centers and 4 community affiliate hospitals, between March 4, 2020 and July 12, 2020. Data for all of these individuals were drawn from the Partners Research Patient Data Registry (RPDR)[7] and the Enterprise Data Warehouse (EDW) and used to generate an i2b2 datamart.[8] Specific data included age, sex, race/ethnicity, as well as inpatient hospitalization status and vital status. Age-adjusted Charlson comorbidity index, a measure of overall burden of illness, was calculated using coded ICD9 and 10 diagnostic codes drawn from the EHR as previously described[9]. Presence of a PCR coronavirus test, and test result, was determined from the enterprise laboratory feed (LOINC:94309-2).

The Partners HealthCare Human Research Committee approved the study protocol. As no participant contact was required in this study based on secondary use of data arising from routine clinical care, the committee waived the requirement for informed consent as detailed by 45 CFR46.116.

### Medication exposure

Medication exposures were defined based upon presence of at least one prescription event or medication order (categorized as RxNorm ingredient) in the electronic health record in the 1 year - 30 days prior presentation to the emergency department (A sensitivity analysis requiring 2 prescription events did not meaningfully change results and is not described further.)

### Study Design and Analysis

Primary analysis sought to examine whether medications were over-represented among a subset of COVID-19 tested individuals. Specifically, to detect medications that might diminish morbidity, we considered individuals hospitalized versus not hospitalized among those testing positive, and secondarily individuals requiring ICU admission or who died during the study period versus not among those hospitalized. (Secondary analyses also considered medication classes, defined by National Drug File - Reference Terminology drug classes, drawn from UMLS[10])

We used multiple logistic regression with outcome as dependent variable and medication ingredient as predictor, with adjustment for age, sex, race, ethnicity, Charlson score, and presence of any diagnosis prior to admission (i.e., availability of prior clinical data). As a hypothesis-generating study, we did not Bonferroni-correct for multiple contrasts, and focused on estimates of odds ratio. To minimize risk of re-identification, in tables, all groups including fewer than 10 individuals are obfuscated. All analyses utilized R4.0.0[11]. Patients and the public were not involved in the design, or conduct, or reporting, or dissemination plans of our research. We do not plan to disseminate the results to study participants or patient organizations.

## Results

Among 7,360 individuals with COVID-19 positive test results, 3,693 (50.2%) were hospitalized in one of 6 hospitals. Table 1 reports sociodemographic and clinical characteristics of individuals testing positive, distinguishing documented inpatient hospitalization or lack of hospitalization.

**Table 1.** Sociodemographic comparisons of study groups. Population includes COVID+ patients presenting to a Mass General Brigham emergency department (ED). Comparison is between patients hospitalized within 7 days of an ED visit.

|  | COVID+ hospitalized<br>patients<br>(N=3693) | COVID+ patients, not<br>hospitalized<br>(N=3667) | Total<br>(N=7360) | p value |
| --- | --- | --- | --- | --- |
| **Hospital type** |  |  |  | < 0.001 |
| Academic medical<br>centers | 2112 (57.2%) | 1861 (50.7%) | 3973 (54.0%) |  |
| Community hospitals | 1581 (42.8%) | 1806 (49.3%) | 3387 (46.0%) |  |
| **Age group** |  |  |  | < 0.001 |
| < 30 | 170 (4.6%) | 852 (23.2%) | 1022 (13.9%) |  |
| 30-39 | 295 (8.0%) | 758 (20.7%) | 1053 (14.3%) |  |
| 40-49 | 409 (11.1%) | 702 (19.1%) | 1111 (15.1%) |  |
| 50-59 | 635 (17.2%) | 664 (18.1%) | 1299 (17.6%) |  |
| 60-69 | 775 (21.0%) | 392 (10.7%) | 1167 (15.9%) |  |
| 70-79 | 658 (17.8%) | 185 (5.0%) | 843 (11.5%) |  |
| 80+ | 751 (20.3%) | 114 (3.1%) | 865 (11.8%) |  |
| **Male gender** | 2024 (54.8%) | 1841 (50.2%) | 3865 (52.5%) | < 0.001 |
| **Race** |  |  |  | < 0.001 |
| Asian | 136 (3.7%) | 137 (3.7%) | 273 (3.7%) |  |
| Black | 624 (16.9%) | 629 (17.2%) | 1253 (17.0%) |  |
| Other | 574 (15.5%) | 795 (21.7%) | 1369 (18.6%) |  |
| Unknown | 397 (10.8%) | 483 (13.2%) | 880 (12.0%) |  |
| White | 1962 (53.1%) | 1623 (44.3%) | 3585 (48.7%) |  |
| **Hispanic ethnicity** | 1005 (27.2%) | 1542 (42.1%) | 2547 (34.6%) | < 0.001 |
| **Charlson comorbidity**<br>**index** |  |  |  | < 0.001 |
| Mean (SD) | 2.588 (3.408) | 0.901 (1.962) | 1.748 (2.908) |  |
| Range | 0-21 | 0-19 | 0-21 |  |
| Median (Q1, Q3) | 1 (0-4) | 0 (0-1) | 0 (0-2) |  |
| **ICU admission** | 962 (26.0%) | 0 (0.0%) | 962 (13.1%) | < 0.001 |
| **Mechanical ventilation** | 829 (22.4%) | 0 (0.0%) | 829 (11.3%) | < 0.001 |
| **Death** | 608 (16.5%) | 55 (1.5%) | 663 (9.0%) | < 0.001 |

Table 2 indicates those medications with the greatest differences in prescribing between hospitalized and not-hospitalized individuals and frequency among test-positive individuals of at least 1%, as well as odds ratios for hospitalization associated with each medication after adjustment for age, sex, race, ethnicity, site, and Charlson score. Among electronically prescribed medications with greatest increase in rank among test-positive individuals not requiring hospitalization, and associated with odds ratios less than 1, were ibuprofen (OR 0.73, 95% CI 0.64-0.84), azelastine (0.39, 95% CI 0.21-0.70), and sumatriptan (0.43, 95% CI 0.25-0.75). Table 2 also includes 3 medicines commonly used in the treatment of substance use disorders or overdose - naloxone, flumazenil, and chlordiazepoxide. (For all medications with frequency greater than 1% among test-positive individuals, see Figure 1 and Supplemental Table 2). The NSAID naproxen also was also associated with diminished probability of hospitalization (OR 0.66, 95% CI 0.50-0.87) (Supplemental Table 2.) Follow-up analyses examined medications by therapeutic category; no individual category was associated with experiment-wide significance for reduction in risk (Table 3 and Figure 2, and Supplemental Table 3).

**Table 2.** Top 20 medication ingredients associated with hospitalization following ED visit among COVID+ patients. Adjusted models were controlled for age at admission, gender, race, ethnicity, Charlson comorbidity index and the presence of a prior diagnosis in the health system.

| RxNorm code | Medication ingredient | NDF-RT medication type | Patients |  | Unadjusted models |  |  |  | Adjusted models |  |  |  |
| --- | --- | --- | --- | --- | --- | --- | --- | --- | --- | --- | --- | --- |
|  |  |  | Patients hospitalized | Patients not hospitalized | OR | 95% CI Low | 95% CI High | p-value | OR | 95% CI Low | 95% CI High | p-value |
| 5640 | Ibuprofen | Musculoskeletal | 626 | 918 | 0.611 | 0.545 | 0.685 | 0.000 | 0.725 | 0.632 | 0.831 | 3.8e-06 |
| 4603 | Furosemide | Cardiovascular | 644 | 149 | 4.987 | 4.146 | 5.998 | 0.000 | 1.653 | 1.321 | 2.069 | 1.1e-05 |
| 5224 | Heparin | Blood products/ modifiers/ volume expanders | 738 | 245 | 3.488 | 2.994 | 4.064 | 0.000 | 1.523 | 1.259 | 1.843 | 1.5e-05 |
| 36387 | Sennosides | Gastrointestinal | 824 | 314 | 3.067 | 2.668 | 3.525 | 0.000 | 1.435 | 1.203 | 1.711 | 5.8e-05 |
| 1596 | Bisacodyl | Gastrointestinal | 522 | 171 | 3.365 | 2.813 | 4.026 | 0.000 | 1.552 | 1.251 | 1.925 | 6.3e-05 |
| 7597 | Nystatin | Antimicrobials | 283 | 72 | 4.144 | 3.186 | 5.390 | 0.000 | 1.838 | 1.357 | 2.488 | 8.3e-05 |
| 5856 | Insulin | Hormones, synthetics, modifiers | 657 | 208 | 3.599 | 3.056 | 4.237 | 0.000 | 1.432 | 1.167 | 1.757 | 0.001 |
| 7242 | Naloxone | Central nervous system | 265 | 261 | 1.009 | 0.845 | 1.205 | 0.923 | 0.700 | 0.566 | 0.865 | 0.001 |
| 121191 | Rituximab | Antineoplastics | 33 | <10 | 6.604 | 2.578 | 16.915 | 0.000 | 5.274 | 1.916 | 14.514 | 0.001 |
| 11124 | Vancomycin | Antimicrobials | 479 | 152 | 3.446 | 2.854 | 4.162 | 0.000 | 1.446 | 1.153 | 1.813 | 0.001 |
| 2356 | Chlordiazepoxide | Central nervous system | 49 | 95 | 0.506 | 0.357 | 0.716 | 0.000 | 0.541 | 0.370 | 0.789 | 0.001 |
| 20481 | Cefepime | Antimicrobials | 294 | 79 | 3.928 | 3.051 | 5.058 | 0.000 | 1.609 | 1.199 | 2.159 | 0.002 |
| 18603 | Azelastine | Nasal and throat agents, topical | 22 | 36 | 0.604 | 0.355 | 1.029 | 0.064 | 0.382 | 0.211 | 0.693 | 0.002 |
| 4457 | Flumazenil | Antidotes, deterrents and poison control | 140 | 121 | 1.155 | 0.901 | 1.479 | 0.255 | 0.639 | 0.483 | 0.847 | 0.002 |
| 48937 | Dexmedetomidine | Central nervous system | 93 | 33 | 2.845 | 1.907 | 4.243 | 0.000 | 2.031 | 1.289 | 3.199 | 0.002 |
| 8570 | Polyvinyl alcohol | Ophthalmic agents | 60 | <10 | 6.713 | 3.329 | 13.534 | 0.000 | 3.368 | 1.540 | 7.368 | 0.002 |
| 37418 | Sumatriptan | Central nervous system | 20 | 61 | 0.322 | 0.194 | 0.534 | 0.000 | 0.424 | 0.243 | 0.739 | 0.002 |
| 1292 | Baclofen | Musculoskeletal | 93 | 38 | 2.467 | 1.687 | 3.608 | 0.000 | 1.932 | 1.258 | 2.968 | 0.003 |
| 67108 | Enoxaparin | Blood products/ modifiers/ volume expanders | 735 | 331 | 2.504 | 2.180 | 2.877 | 0.000 | 1.299 | 1.094 | 1.543 | 0.003 |

**Figure 1.**
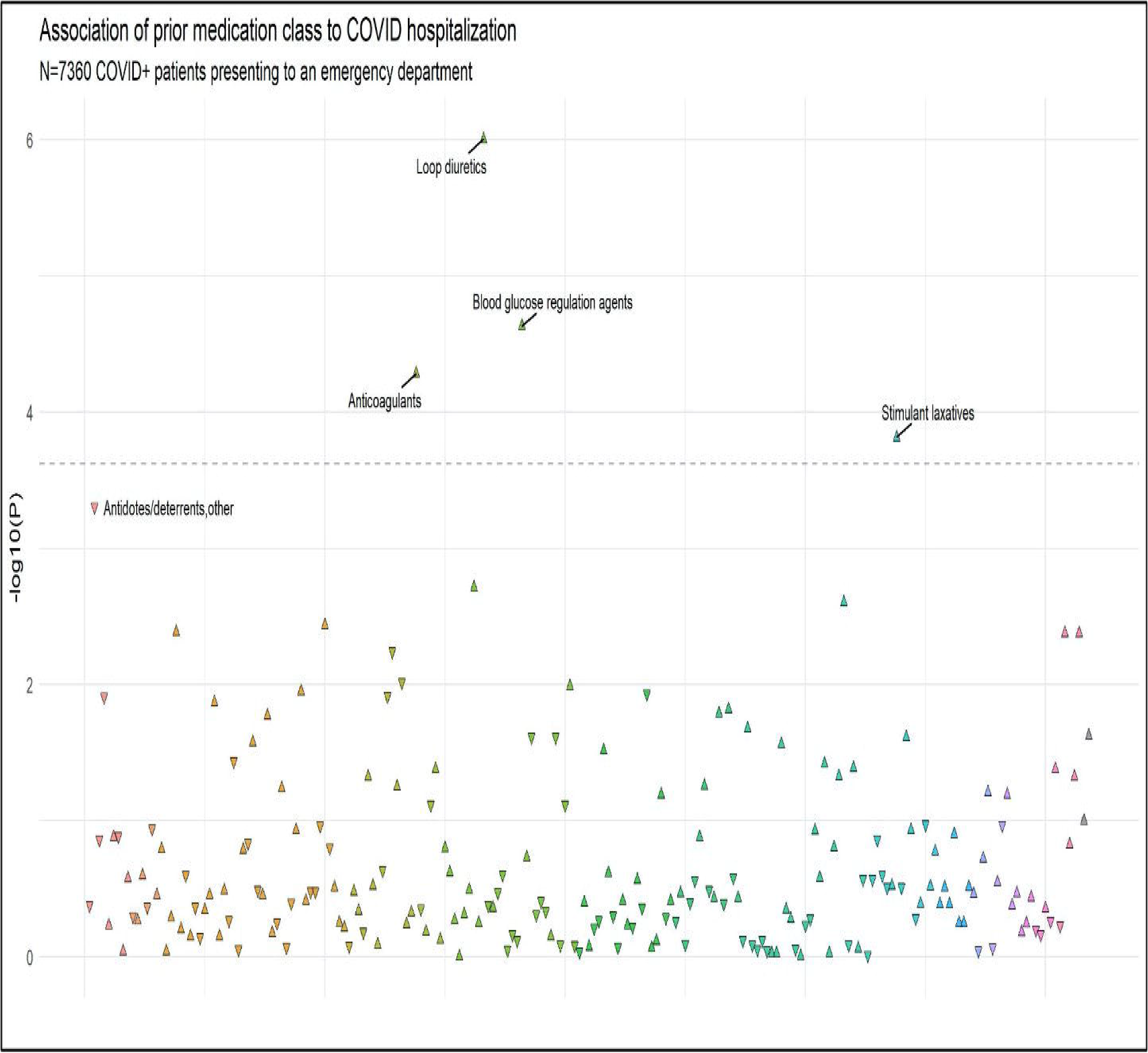
Manhattan plot of prior medications associated with hospitalization among COVID+ patients presenting to an emergency room. Each triangle represents a single medication ingredient. The y-axis corresponds to the negative log 10 of the adjusted model p-value. Models were adjusted for age at admission, gender, race, ethnicity, Charlson comorbidity index and the presence of a prior diagnosis in the health system. Upward facing triangles represent medications conveying increased risk (OR>1) and downward facing triangle indicate decreased risk (OR<1) in adjusted models. A total of 442 medication ingredients that were prescribed to at least 20 hospitalized patients were evaluated. The dashed line is the Bonferroni level of significance. Medication ingredients above and just below the Bonferroni line are labeled.

**Table 3.** Top 20 medication classes associated with hospitalization following ED visit among COVID+ patients. Adjusted models were controlled for age at admission, gender, race, ethnicity, Charlson comorbidity index and the presence of a prior diagnosis in the health system.

| NDF-RT code | NDF-RT medication class | NDF-RT medication type | Patients hospitalized | Patients not hospitalized | Unadjusted models |  |  |  | Adjusted models |  |  |  |
| --- | --- | --- | --- | --- | --- | --- | --- | --- | --- | --- | --- | --- |
|  |  |  |  |  | OR | 95% CI Low | 95% CI High | p-value | OR | 95% CI Low | 95% CI High | p-value |
| N0000029127 | Loop diuretics | Cardiovascular medications | 648 | 129 | 5.361 | 4.414 | 6.512 | 0.000 | 1.798 | 1.421 | 2.274 | 4.2e-07 |
| N0000029183 | Blood glucose regulation agents | Cardiovascular medications | 881 | 318 | 3.031 | 2.639 | 3.480 | 0.000 | 1.442 | 1.217 | 1.709 | 1.2e-06 |
| N0000029110 | Anticoagulants | Blood products/modifiers /volume expanders | 1159 | 435 | 3.129 | 2.767 | 3.538 | 0.000 | 1.397 | 1.188 | 1.643 | 2.0e-05 |
| N0000029392 | Stimulant laxatives | Gastrointestinal medications | 864 | 312 | 3.031 | 2.637 | 3.484 | 0.000 | 1.406 | 1.178 | 1.676 | 2.8e-05 |
| N0000029070 | Antidotes/deterrents, other | Antidotes, deterrents and poison control | 300 | 247 | 1.103 | 0.927 | 1.314 | 0.269 | 0.696 | 0.567 | 0.853 | 1.0e-04 |
| N0000029125 | Diuretics | Cardiovascular medications | 895 | 281 | 3.553 | 3.079 | 4.102 | 0.000 | 1.322 | 1.108 | 1.577 | 0.002 |
| N0000029185 | Oral hypoglycemic agents, oral | Diagnostic agents | 560 | 226 | 2.493 | 2.120 | 2.932 | 0.000 | 1.332 | 1.106 | 1.603 | 0.002 |
| N0000029554 | Cephalosporin 4th generation | Antimicrobials | 278 | 71 | 3.771 | 2.897 | 4.909 | 0.000 | 1.573 | 1.159 | 2.135 | 0.004 |
| N0000029079 | Cephalosporin 3rd generation | Antimicrobials | 582 | 207 | 2.871 | 2.432 | 3.390 | 0.000 | 1.347 | 1.099 | 1.651 | 0.004 |
| N0000029273 | Vitamins, combinations | Vitamins | 64 | 24 | 2.471 | 1.542 | 3.959 | 0.000 | 2.234 | 1.289 | 3.874 | 0.004 |
| N0000029466 | Multivitamins with minerals | Vitamins | 64 | 24 | 2.471 | 1.542 | 3.959 | 0.000 | 2.234 | 1.289 | 3.874 | 0.004 |
| N0000029241 | Bronchodilators, sympathomimetic, oral | Autonomic medications | 798 | 532 | 1.473 | 1.304 | 1.663 | 0.000 | 0.799 | 0.682 | 0.937 | 0.006 |
| N0000029301 | Antivertigo agents | Autonomic medications | 116 | 101 | 1.073 | 0.819 | 1.406 | 0.609 | 0.657 | 0.479 | 0.903 | 0.010 |
| N0000029370 | Emollients | Cardiovascular medications | 165 | 50 | 3.062 | 2.228 | 4.207 | 0.000 | 1.622 | 1.121 | 2.346 | 0.010 |
| N0000029366 | Beta-lactams antimicrobials, other | Antimicrobials | 72 | 12 | 5.593 | 3.030 | 10.321 | 0.000 | 2.351 | 1.215 | 4.551 | 0.011 |
| N0000029300 | Antimigraine agents | Central nervous system medications | 26 | 80 | 0.293 | 0.187 | 0.457 | 0.000 | 0.530 | 0.324 | 0.869 | 0.012 |
| N0000029240 | Bronchodilators,<br>sympathomimetic,<br>inhalation | Autonomic medications | 800 | 542 | 1.445 | 1.280 | 1.631 | 0.000 | 0.818 | 0.699 | 0.957 | 0.012 |
| N0000029134 | Opioid antagonist<br>analgesics | Antidotes, deterrents<br>and poison control | 271 | 235 | 1.046 | 0.873 | 1.253 | 0.625 | 0.762 | 0.616 | 0.943 | 0.012 |
| N0000029090 | Anti-infectives, other | Antimicrobials | 683 | 308 | 2.277 | 1.972 | 2.629 | 0.000 | 1.254 | 1.048 | 1.500 | 0.013 |
| N0000029358 | Electrolytes/minerals | Dental and oral agents,<br>topical | 992 | 482 | 2.230 | 1.976 | 2.518 | 0.000 | 1.218 | 1.039 | 1.428 | 0.015 |

**Figure 2.**
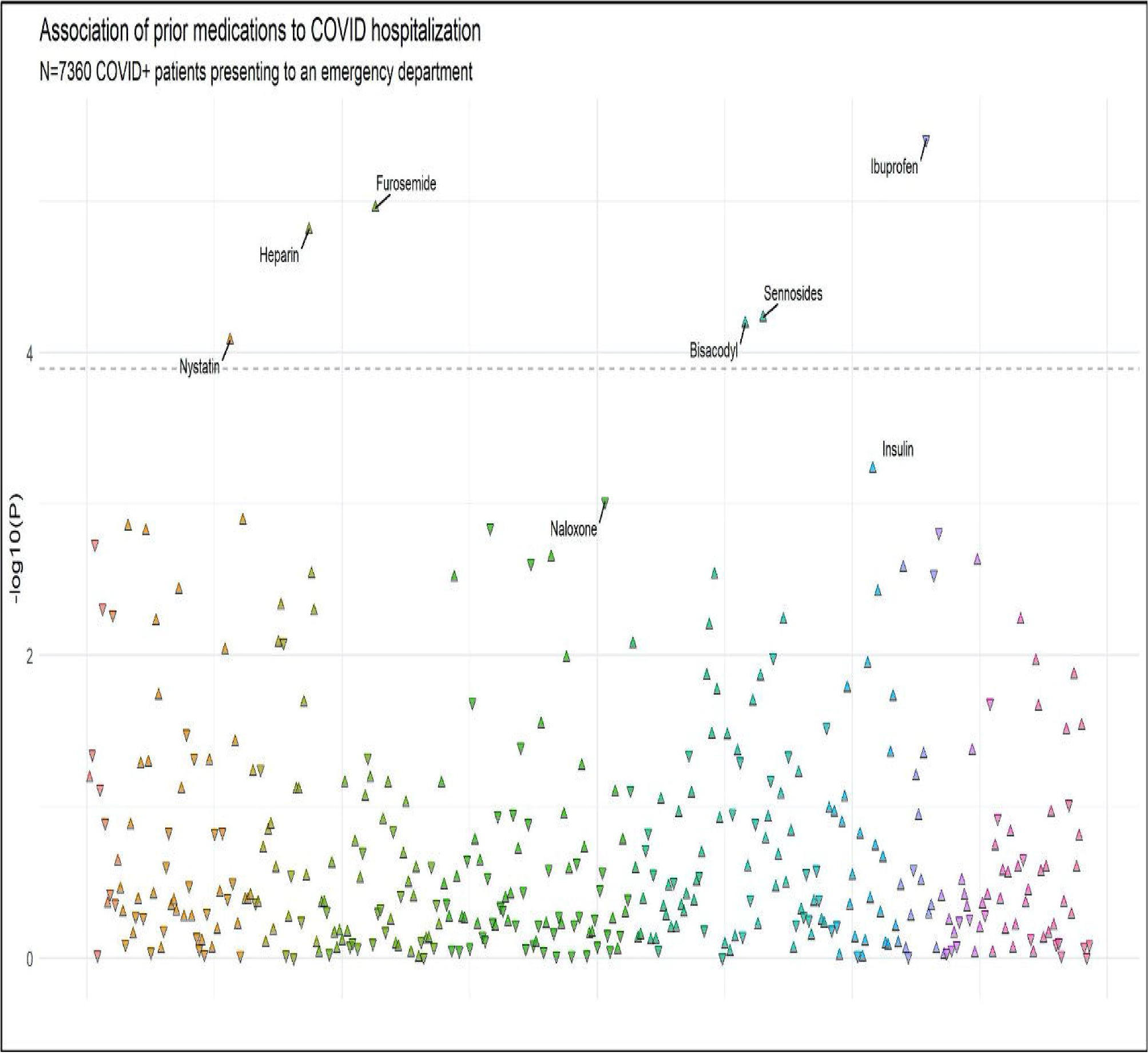
Manhattan plot of prior medication classes associated with hospitalization among COVID+ patients presenting to an emergency room. Each triangle represents a medication class. The y-axis corresponds to the negative log 10 of the adjusted model p-value. Models were adjusted for age at admission, gender, race, ethnicity, Charlson comorbidity index and the presence of a prior diagnosis in the health system. Upward facing triangles represent medication classes conveying increased risk (OR>1) and downward facing triangle indicate decreased risk (OR<1) in adjusted models. A total of 204 medication classes that were prescribed to at least 20 hospitalized patients were evaluated. The dashed line is the Bonferroni level of significance. Medication classes above and just below the Bonferroni line are labeled.

Among individuals who were hospitalized, 962 (26.0%) were admitted to the ICU and 608 (16.5%) died; features of this cohort are reported in Supplemental Data. Adjusted odds ratios for ICU admission and death associated with each medication are also listed; ibuprofen was significantly associated with decreased odds of requiring ICU (OR 0.70, 95% CI 0.56-0.86) and mortality (0.73, 95% CI 0.56-0.96).

## Discussion

This preliminary electronic health records study examined prehospitalization medications associated with differential outcomes among 7,360 individuals who tested positive for COVID-19 and were seen in the emergency department or hospitalized in one of 6 Boston-area hospitals between 3/5/2020 and 7/12/2020. While not the primary intention of this analysis, we identify markers of prior treatment in long-term care or skilled nursing facilities (e.g., bowel regimens) and markers of known comorbidities associated with poorer outcomes (e.g., renal disease) as associating with greater risk of hospitalization, suggesting assay sensitivity at least for risk-increasing medications. We also identify multiple medications (e.g., those used for substance use disorders or overdoses) associated with lower risk of hospitalization, suggesting that these may be high-risk individuals who test positive for COVID-19 but are not necessarily seen in the emergency department for COVID-19 (e.g., individuals who misuse drugs and alcohol). Both of these examples of probable confounding underscore the extreme caution required in interpreting nonrandomized clinical data.

However, we do identify individual candidate medications that, if further supported in other cohorts, might be screened for repositioning as COVID-19 treatment or for prevention of morbidity. Most notably, two nonsteroidal anti-inflammatory drugs, ibuprofen and naproxen, appear to be enriched among patients not requiring hospitalization, and show qualitatively similar enrichment among patients not requiring mechanical ventilation during hospitalization. We emphasize that we cannot exclude confounding in this context as well; dysregulated immunity in autoimmune disease could associate with differential severity. However, it also seems plausible that suppression of an inflammatory response may diminish the immune consequences of COVID-19 infection that may contribute to pathogenesis[12], especially in light of recent reports associating cytokine level with clinical outcome[13]. Initial case reports of greater morbidity among NSAID-treated patients, postulated to arise from increases in the cellular COVID-19 receptor ACE-2, have subsequently been questioned[14].

Multiple FDA-approved medications have been suggested in in vitro studies to diminish coronavirus infection in general, or COVID19 in particular. For example, prior work identified multiple neurotransmitter inhibitors, including chlorpromazine, as inhibiting coronavirus infection in kidney epithelial cells[2]; a similar effort identified 4 compounds (chloroquine, chlorpromazine, loperamide, and lopinavir) as inhibiting coronavirus replication [15]. A recent in silico study examining affinity for the key COVID-19 protease[3] identified ziprasidone as a potential covalently-binding inhibitor. Our analysis does not provide further support for chloroquine, and is not informative regarding the others However, as additional in vitro data emerges, our results may be applied to provide orthogonal evidence to support further investigation.

We emphasize the preliminary nature of these findings, which will require replication and extension in other data sets prior to clinical investigation. Two key limitations must be emphasized. First, as in any nonrandomized study, the risk for bias - and particularly for confounding by indication - is high. That is, other differences between the groups being compared, such as differences in comorbidity, may be proxied by the medication exposure; causal relationships cannot be inferred. Indeed, the association with greater adverse outcomes with renal disease and diabetes treatments, and with bowel regimens commonly seen in skilled nursing facilities, likely represents such residual confounding.

Second, the power afforded by this cohort, despite incorporating a network of hospitals, is modest. In particular, absence of effect does not preclude potential efficacy, particularly as our approach will exclude rarer medications, such as those typically prescribed for time-limited periods such as short-term antibiotics.

Further limitations include the reliance on medications electronically prescribed, rather than filled, in the 12 months prior to hospitalization. The lag in availability of current medication data precludes characterization of current medications on admission. This lag, as well as the possibility that medications may be discontinued or not filled, would tend to bias results toward the null hypothesis, by introducing additional heterogeneity. While sensitivity analysis requiring multiple prescriptions for definition of exposure did not meaningfully change results, further studies with more precise exposure characterization will be critical once larger cohorts are available.

Despite these limitations, medication repositioning represents an appealing strategy for responding to COVID-19 because of the rapidity with which promising interventions can be transitioned to clinical trials, and potentially to clinical application [4]. As the safety profile of these medications is well-understood, trials can focus on detection of short-term efficacy, without requiring the longer-term safety data typically required for FDA registration[4,6]. A particularly powerful strategy may integrate clinical informatics and cellular/translational data, as such data become available. At minimum, we hope these results will spur others to pursue similar investigations, yielding the larger, richer data sets required for detection of more modest effects and control of confounding.

## Data Availability

Institutional review board approval does not allow sharing of identifiable data outside of the health system, but all analyses are provided as Supplemental Materials.

## Contributor and Guarantor Information

No funding source contributed to any aspect of study design, data collection, data analysis, or data interpretation. Dr. Perlis had full access to all the data in the study and accepts full responsibility for the work. Dr. Perlis conceived the research question, planned the analysis, and drafted and revised the manuscript. Mr. Castro generated the data set, conducted analyses, and revised the manuscript. Dr. Ross and Dr. McBride contributed to the analytic plan and assisted in manuscript revision. All authors shared the final responsibility for the decision to submit for publication. Dr. Perlis attests that all listed authors meet authorship criteria and that no others meeting the criteria have been omitted. Dr. Perlis also attests that the manuscript is an honest, accurate, and transparent account of the study being reported; that no important aspects of the study have been omitted and that any discrepancies from the study as originally planned have been explained.

The Corresponding Author (Dr. Perlis) has the right to grant on behalf of all authors and does grant on behalf of all authors, a worldwide license to the Publishers and its licensees in perpetuity, in all forms, formats and media (whether known now or created in the future), to i) publish, reproduce, distribute, display and store the Contribution, ii) translate the Contribution into other languages, create adaptations, reprints, include within collections and create summaries, extracts and/or, abstracts of the Contribution, iii) create any other derivative work(s) based on the Contribution, iv) to exploit all subsidiary rights in the Contribution, v) the inclusion of electronic links from the Contribution to third party material where-ever it may be located; and, vi) license any third party to do any or all of the above.

